# The Lost Glucose Sink: Association of Sarcopenia Risk with Disproportionate Postprandial Glycemic Excursions in Older Adults with Type 2 Diabetes Mellitus

**DOI:** 10.64898/2026.10.05.26364797

**Authors:** Debankur Chakraborty

**Affiliations:** R.G.Kar Medical College Kolkata

**Keywords:** sarcopenia, SARC F, type 2 diabetes mellitus, postprandial hyperglycemia, skeletal muscle, older adults

## Abstract

**Background:** Many adults with type 2 diabetes mellitus (T2DM) in India have a “thin fat” body type, with low skeletal muscle mass for their body mass index. Muscle is the main site of glucose disposal after a meal. Fasting blood sugar (FBS) mostly reflects glucose output from the liver, whereas postprandial blood sugar (PPBS) depends more on insulin driven glucose uptake by muscle.

**Objective:** To show how the association between sarcopenia risk and postprandial glycemic excursion would be tested in older adults with T2DM and controlled fasting glucose.

**Methods:** A cross sectional design is planned for adults aged 50 years or more with T2DM, FBS of 90–130 mg/dL and a stable oral regimen. Sarcopenia risk is measured with the SARC F questionnaire (score ≥4 = high risk). Glycemic excursion (ΔBG) is PPBS minus FBS. The method was applied to a dataset of 300 patients collected over an OPD setting. Groups were compared with Welch‚s t test, the relationship between SARC F score and ΔBG was tested with Pearson‚s correlation, and multiple linear regression adjusted for age, diabetes duration, FBS and drug regimen.

**Results:** In the patient dataset (mean age 67.6 ± 6.8 years), 64 patients (21.3%) had a high risk SARC F score. Mean ΔBG was 119.4 ± 29.1 mg/dL in the high risk group and 74.8 ± 22.8 mg/dL in the low risk group (difference 44.6 mg/dL, 95% CI 36.8–52.5; p<0.0001). SARC F score and ΔBG were positively correlated (r = 0.777, p<0.0001). After adjustment, each 1 point rise in SARC F score was linked to a 10.2 mg/dL higher ΔBG (95% CI 8.6–11.7). The high risk group was about 6 years older, and age and diabetes duration explained part of the crude difference.

**Conclusion:** The analysis plan can detect and quantify a link between SARC F score and post meal glucose rise while accounting for age, diabetes duration, FBS and drug regimen. Clinical treatment algorithms must shift from exclusively targeting insulin secretion to restoring the mechanical glucose sink through resistance training and age appropriate pharmacological adjustments

## 1. Introduction

Type 2 diabetes mellitus (T2DM) has a distinct pattern in the Indian subcontinent. Compared to white Europeans of the same body mass index, Asian Indians tend to carry more visceral fat and less lean skeletal muscle, a pattern often called the “thin fat” or “Asian Indian” phenotype [1,2].

Fasting blood sugar (FBS) mainly reflects the glucose made by the liver overnight. After a meal, the picture changes: skeletal muscle takes up most of the glucose that insulin clears from the blood, with estimates of up to about 80% [3,4]. Lower relative muscle mass has also been linked to higher insulin resistance [5]. Muscle can therefore be seen as a “glucose sink”, and a smaller sink may mean a bigger rise in blood sugar after eating.

Sarcopenia is the progressive loss of skeletal muscle mass, strength and function that comes with age [6 8]. Diabetes speeds up this loss, and older people with T2DM have weaker and poorer muscle than those without diabetes [9,10]. In people whose HbA1c is close to target, a large share of the total glucose burden comes from the post meal period [11].

In busy Indian outpatient clinics, doctors often see elderly patients whose fasting sugar is well controlled but whose postprandial sugar remains high. The usual response is to raise the dose of an insulin secretagogue such as a sulfonylurea. We suspected that some of these isolated post meal spikes are not caused only by failing pancreatic beta cells, but also by unrecognized loss of muscle function. This study aims to correlate functional muscle deficiency with increased postprandial glycemic excursions using the rapid SARC F screening tool

## 2. Materials and Methods

### 2.1 Study Design, Setting and Data Status

The study is a single centre, cross sectional observational study of patients attending the Medicine outpatient department (OPD) of R.G. Kar Medical College and Hospital, Kolkata. Patients on routine follow up with recent FBS and PPBS reports were included, and no blood tests were done for the study. Data collection was done from these patients over a period of two months. For each patient, the age, diabetes duration, oral drug regimen, FBS, PPBS and SARC F score were be recorded. Reporting followed the STROBE statement for observational studies [13].

### 2.2 Eligibility Criteria

To study muscle function while limiting the effect of other factors, strict eligibility criteria was used.

#### Inclusion criteria

- **Confirmed T2DM:** diabetes duration of 3 years or more.
- **Age:** adults aged 50 years or more, to cover the period when age related and diabetes related sarcopenia begins.
- **Controlled fasting glucose:** FBS between 90 and 130 mg/dL on the report used for the study. This ensures that the postprandial rise is a true post meal excursion and not a carry over from poorly controlled fasting glucose.ss
- **Stable treatment:** a stable regimen of oral hypoglycemic agents (OHAs), for example metformin, DPP 4 inhibitors or sulfonylureas, for at least 3 months.

#### Exclusion criteria

- **Insulin therapy:** current use of basal or prandial insulin, because it bypasses the body‚s own glucose clearance.
- **Advanced kidney or liver disease:** chronic kidney disease with eGFR <45 mL/min/1.73 m^2^, or active liver disease, as both change how oral drugs act.
- **Walking difficulty unrelated to sarcopenia:** for example, a recent fracture, severe knee osteoarthritis or hemiparesis after stroke.
- **Acute illness:** any active infection or acute inflammatory illness in the previous 14 days, which can cause temporary stress hyperglycemia.

### 2.3 Assessment of Sarcopenia Risk

Sarcopenia risk was assessed with the SARC F questionnaire, a simple five item screening tool [12]. It asks about Strength, Assistance in walking, Rising from a chair, Climbing stairs and Falls. Each item scores 0 to 2, so the total ranges from 0 to 10. A score of 4 or more indicates high risk of sarcopenia and related functional decline, and the tool has been tested in community dwelling older adults [14].

### 2.4 Glycemic Measures and Drug Regimen

FBS and PPBS values were taken from each patient‚s most recent report. Glycemic excursion (ΔBG) is defined as PPBS minus FBS, in mg/dL. Each regimen was grouped by the drug added to metformin: metformin alone (n = 83), metformin plus a sulfonylurea (glimepiride; n = 96) and metformin plus a DPP 4 inhibitor (sitagliptin, teneligliptin or vildagliptin; n = 121).

### 2.5 Statistical Analysis

Continuous variables are shown as mean ± standard deviation (SD) and categorical variables as number (percentage). Patients were divided into a low risk group (SARC F <4) and a high risk group (SARC F ≥4). A two sided p value below 0.05 was taken as statistically significant. Analyses were done in Python (SciPy 1.17 and statsmodels 0.15). Normality was checked with the Shapiro–Wilk test and equality of variances with Levene‚s test. The main statistical measures are explained below in simple terms.

#### Independent samples t test

This test compares the average of one measurement between two separate groups; here, the mean ΔBG of the low risk and high risk patients. It works out how large the gap between the two group mean is, compared with the normal scatter of values within the groups. This ratio is the t value: the gap divided by its standard error. A larger t value (positive or negative) means the gap is large compared with the scatter, and so is less likely to be due to chance. The t value is then converted into a p value using the t distribution and the degrees of freedom (a number that depends on the sample size). The usual (Student‚s) version assumes that the spread of values is the same in both groups.

Welch‚s version does not need this assumption and gives almost the same answer when the spreads are equal, so we used Welch‚s t test for all group comparisons [15]. The test also assumes independent observations and roughly normal data; with groups of this size it is fairly robust to mild departures from normality. As a check, we repeated the main comparisons with the Mann–Whitney U test, which uses ranks and does not need normal data.

#### Pearson‚s correlation coefficient (r)

This measures how closely two continuous measurements follow a straight line relationship; here, SARC F score and ΔBG. The value of r ranges from −1 to +1. The sign shows the direction: a positive r means ΔBG tends to rise as the SARC F score rises. The size shows the strength: 0 means no linear relationship and 1 means a perfect one. As a rough guide, r values of 0.40–0.69 are called moderate and 0.70–0.89 strong [16]. Squaring r (r^2^) tells us the share of the variation in one measure that is accounted for by the other. A correlation shows that two measures move together; it does not show that one causes the other. Because SARC F scores are whole numbers from 0 to 10, we also calculated Spearman‚s rank correlation (^ρ^), which does not assume a straight line relationship.

#### p value

The p value is the probability of getting a result at least as extreme as the one observed if there were truly no difference between the groups (or no relationship between the two measures). A small p value (below 0.05) means the data are hard to explain by chance alone, so we reject the idea of “no difference”. The p value does not tell us how large or how important an effect is, and it is not the probability that our hypothesis is true [17]. For this reason, we report the size of each effect (mean differences with 95% confidence intervals, and r) together with its p value. Very small p values are reported as p<0.0001.

#### Adjusted and stratified analyses

Because older age and longer diabetes can both affect muscle and glucose, we fitted a multiple linear regression model with ΔBG as the outcome and SARC F score, age, diabetes duration, FBS and drug regimen as predictors. The coefficient for SARC F gives the average change in ΔBG for each 1 point rise in score, with the other variables held constant. The spread of the residuals was uneven (Breusch–Pagan p<0.001), so we used heteroscedasticity robust (HC3) standard errors. We also compared ΔBG between risk groups within three age bands (< 65, 65–75 and >75 years), and compared ΔBG and risk group across the three drug regimens.

## 3. Results

### 3.1 Patient Characteristics and Baseline Glycemia

The dataset had 300 records, all meeting the inclusion criteria. Of these, 236 (78.7%) were at low risk and 64 (21.3%) were at high risk of sarcopenia. The mean age was 67.6 ± 6.8 years (range 50–85) and the mean diabetes duration was 11.1 ± 5.5 years. The high risk group was about 6 years older (72.2 vs 66.3 years, p<0.0001) and had a slightly longer diabetes duration (12.5 vs 10.7 years, p = 0.035). FBS was similar in the two groups (113.4 vs 110.9 mg/dL, p = 0.095) (Table 1). By design, fasting glucose was well controlled (mean FBS 111.5 ± 10.7 mg/dL). Post meal glucose was higher, with a mean PPBS of 195.7 ± 33.9 mg/dL (range 122–316) and a mean excursion (ΔBG) of 84.3 ± 30.4 mg/dL.

**Table 1.**
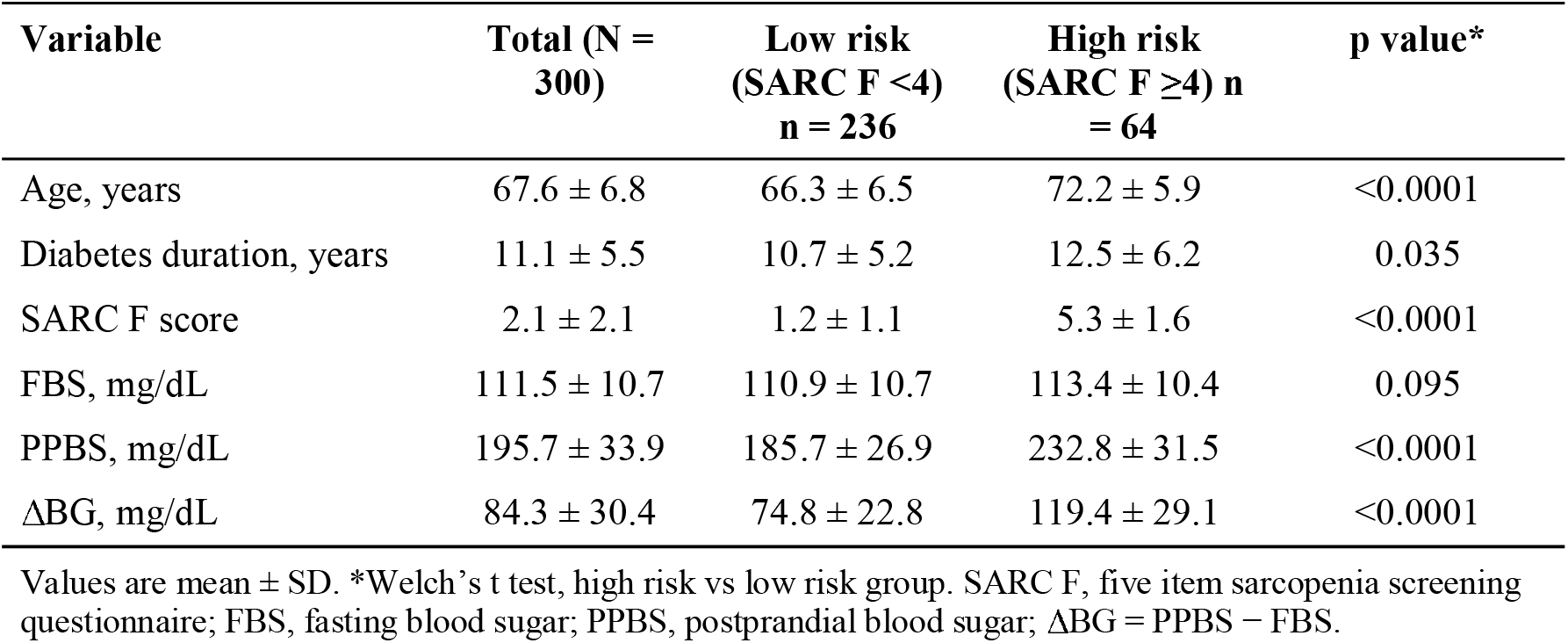
Characteristics and glycemic measures, by sarcopenia risk group.

| Variable | Total (N = 300) | Low risk (SARC F <4) n = 236 | High risk (SARC F ≥4) n = 64 | p value* |
| --- | --- | --- | --- | --- |
| Age, years | 67.6 ± 6.8 | 66.3 ± 6.5 | 72.2 ± 5.9 | <0.0001 |
| Diabetes duration, years | 11.1 ± 5.5 | 10.7 ± 5.2 | 12.5 ± 6.2 | 0.035 |
| SARC F score | 2.1 ± 2.1 | 1.2 ± 1.1 | 5.3 ± 1.6 | <0.0001 |
| FBS, mg/dL | 111.5 ± 10.7 | 110.9 ± 10.7 | 113.4 ± 10.4 | 0.095 |
| PPBS, mg/dL | 195.7 ± 33.9 | 185.7 ± 26.9 | 232.8 ± 31.5 | <0.0001 |
| ΔBG, mg/dL | 84.3 ± 30.4 | 74.8 ± 22.8 | 119.4 ± 29.1 | <0.0001 |

Values are mean ± SD. \*Welch's t test, high risk vs low risk group. SARC F, five item sarcopenia screening questionnaire; FBS, fasting blood sugar; PPBS, postprandial blood sugar; ΔBG = PPBS – FBS.

### 3.2 Association Between Sarcopenia Risk and Glycemic Excursion

High risk patients (SARC F ≥4) had a larger postprandial excursion than low risk patients (119.4 ± 29.1 vs 74.8 ± 22.8 mg/dL), about 1.6 times higher. The mean difference was 44.6 mg/dL (95% CI 36.8–52.5; Welch t = 11.4, df = 85; p<0.0001; Cohen‚s d = 1.8). The median ΔBG was 121.0 mg/dL (IQR 97.3–140.3) in the high risk group and 74.0 mg/dL (IQR 58.0–89.3) in the low risk group (Mann–Whitney p<0.0001). The groups differed in PPBS by 47.1 mg/dL (95% CI 38.6–55.7; p<0.0001) but not clearly in FBS (difference 2.5 mg/dL, 95% CI −0.4 to 5.4; p = 0.095) (Figure 1). Levene‚s test showed unequal variances for ΔBG (p = 0.013) and PPBS (p = 0.040), which supports the use of Welch‚s test.

**Figure 1.**
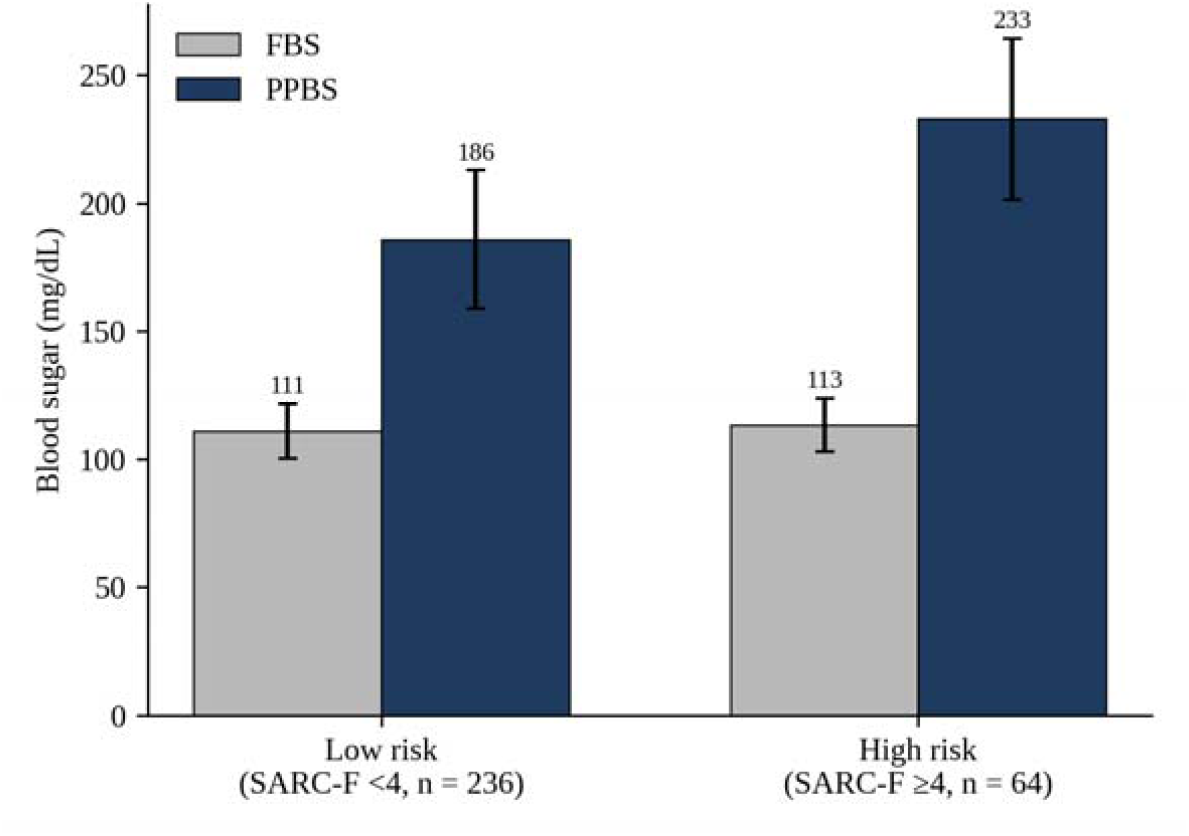
Mean fasting (FBS) and postprandial (PPBS) blood sugar by sarcopenia risk group.

SARC F score had a strong positive correlation with ΔBG (Pearson‚s r = 0.777, 95% CI 0.727–0.818, p<0.0001; Spearman‚s ^ρ^ = 0.751, p<0.0001; Figure 2). This means that about 60% of the variation in glycemic excursion (r^2^ = 0.60) moves in line with the SARC F score. The relationship was also seen inside each group (r = 0.64 in the low risk and r = 0.57 in the high risk group), and mean ΔBG rose steadily from 57 mg/dL at a score of 0 to 141 mg/dL at a score of 7. Few patients had scores of 8 or more (n = 5), so those means are unstable. SARC F score was related to age (r = 0.46, p<0.0001) and to diabetes duration (r = 0.20, p<0.001), but not to FBS (r = 0.09, p = 0.13).

**Figure 2.**
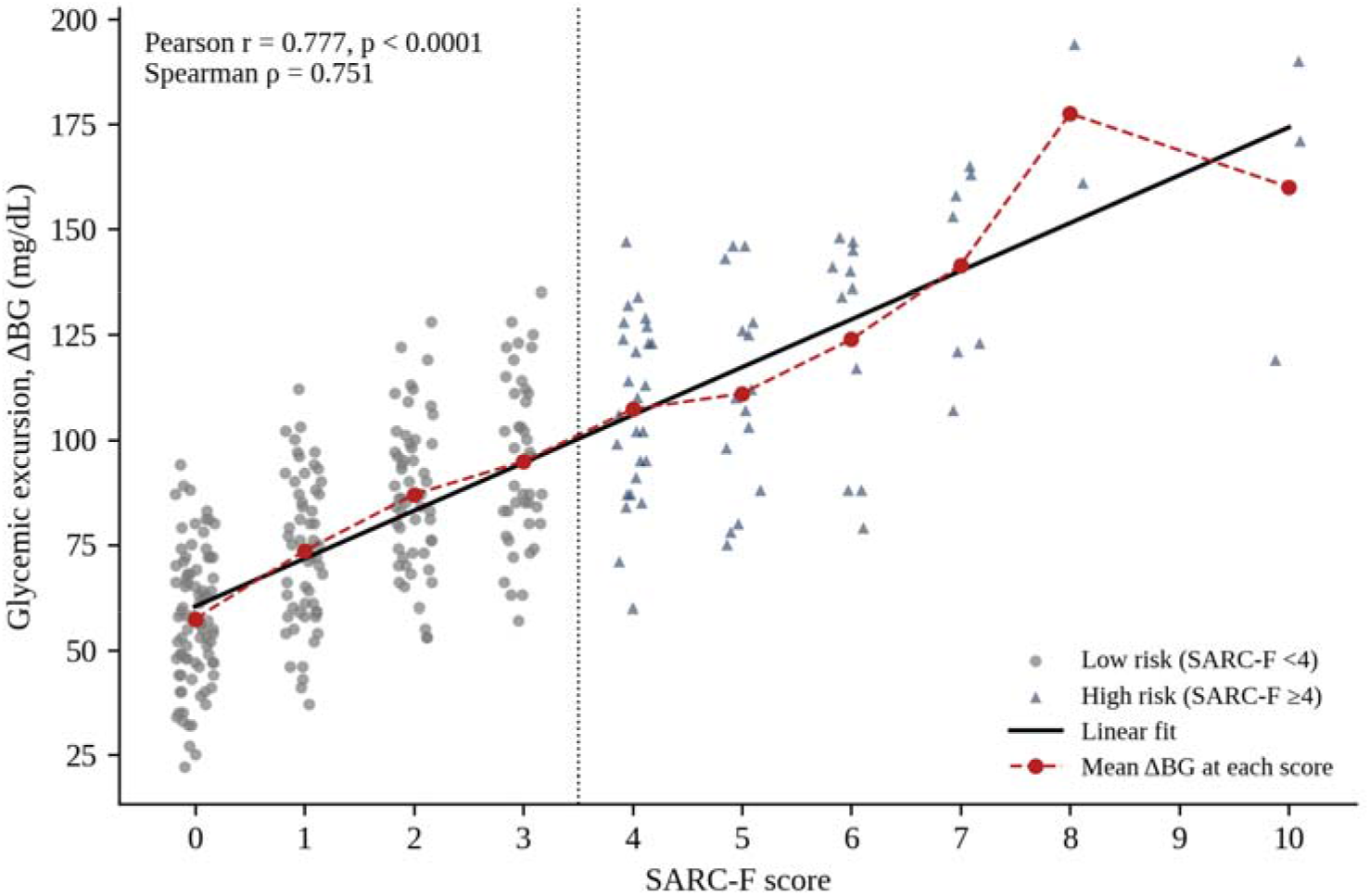
Relationship between SARC F score and glycemic excursion (ΔBG). Each point is one record. The dotted line marks the cut off between low and high risk.

### 3.3 Adjusted and Stratified Analyses

Because the high risk group was older and had a longer diabetes duration, we adjusted for these factors. In the regression model, each 1 point rise in SARC F score was linked to a 10.2 mg/dL higher ΔBG (95% CI 8.6–11.7; p<0.0001) after adjusting for age, diabetes duration, FBS and drug regimen. The unadjusted value was 11.4 mg/dL (95% CI 10.1–12.7). Each extra year of diabetes added 1.4 mg/dL (95% CI 1.1–1.8) and each 1 mg/dL of FBS added 0.3 mg/dL (95% CI 0.1–0.5), whereas age was not significant in this model (p = 0.20). Drug regimen was not a significant predictor. The model explained about 68% of the variation in ΔBG (R^2^ = 0.68). When risk group was used instead of the score, the adjusted difference between high risk and low risk patients was 35.1 mg/dL (95% CI 27.3–43.0), smaller than the crude difference of 44.6 mg/dL. Age and diabetes duration therefore explain part of the crude gap, but not all of it.

The gap in ΔBG between the risk groups was seen in all three age bands (Table 2), but high risk became much more common with age (5.4% below 65 years, 24.0% at 65–75 years and 53.1% above 75 years). The numbers were small in some cells (5 high risk patients below 65 years and 15 low risk patients above 75 years), so these estimates are imprecise.

**Table 2.** Glycemic excursion (ΔBG) by sarcopenia risk group within age bands.

| Age band | n | Low risk: n, ΔBG | High risk: n, ΔBG | Difference (95% CI) |
| --- | --- | --- | --- | --- |
| <65 years | 93 | 88; 68.2 ± 22.2 | 5; 132.2 ± 23.6 | 64.0 (35.0–93.0) |
| 65–75 years | 175 | 133; 77.5 ± 21.9 | 42; 117.6 ± 31.0 | 40.1 (29.8–50.4) |
| >75 years | 32 | 15; 89.3 ± 23.3 | 17; 120.1 ± 26.1 | 30.9 (13.0–48.7) |

ΔBG in mg/dL, mean ± SD. Difference = high risk minus low risk, with 95% CI (Welch). $p < 0.01$ in every age band.

The three drug regimens were similar in ΔBG, in the share of high risk patients, and in age and FBS (Table 3). Mean ΔBG did not differ across regimens (ANOVA p = 0.66) and the share of high risk patients did not differ either (chi square p = 0.38).

**Table 3.** Glycemic excursion and sarcopenia risk by drug regimen.

| Regimen | n | ΔBG, mg/dL | High risk, n (%) |
| --- | --- | --- | --- |
| Metformin alone | 83 | 86.6 ± 35.1 | 15 (18.1) |
| Metformin + sulfonylurea | 96 | 84.3 ± 28.1 | 25 (26.0) |
| Metformin + DPP 4 inhibitor | 121 | 82.7 ± 28.7 | 24 (19.8) |

ΔBG is mean ± SD.

## 4. Discussion

In this dataset, higher sarcopenia risk went hand in hand with a larger rise in blood sugar after meals. This was seen even though all patients had controlled fasting glucose and the same kind of stable oral regimen. Fasting values were similar in both groups, while PPBS differed by about 47 mg/dL. This pattern suggests that the difference lies in how glucose is handled after a meal, and not in basal glucose output. It fits with the role of muscle as the main site of insulin mediated glucose uptake [4,5], and with earlier work linking diabetes and sarcopenia [9,10,18]. The relationship also appeared as a steady rise across SARC F scores and inside each risk group, and not only as a gap between two groups.

The high risk group was older and had a longer diabetes duration, and both were linked to larger excursions. Adjustment reduced the group difference from 44.6 to 35.1 mg/dL, but a clear difference remained. When muscle mass decreases, there is less tissue to take up glucose after a meal. Also, fat builds up inside muscle and low grade inflammation may reduce its response to insulin further [10,18]. The link may also run both ways: poor glycemic control and insulin resistance can themselves speed up muscle loss [18]. A cross sectional design cannot separate these possibilities, and factors such as sex, body mass index, diet and physical activity would also need to be recorded.

Practically, SARC F takes only a few minutes and needs no equipment. Therefore, a quick SARC F score could lead the doctor to think about muscle function when an older patient comes with a high PPBS with a controlled FBS, before simply increasing the dose of a secretagogue.

### 4.1 Clinical Implications: Age and Frailty Stratified Approach

The data definitively shows that older Indian patients experiencing massive PPBS spikes alongside normal FBS levels are predominantly those suffering from functional skeletal muscle decline. This establishes muscle mass not just as a structural asset, but as a critical endocrine organ. Currently, the standard clinical reflex for elevated PPBS in India is the escalation of oral secretagogues. Based on our findings, we propose the following Age and Frailty Stratified Treatment Algorithm to mitigate hypoglycemia and restore the physiological glucose sink.

#### Age below 65 years (pre sarcopenic, low frailty)

Muscle mass is usually still preserved, but insulin resistance is growing. Metformin remains the cornerstone of treatment. If kidney function allows, and especially when cardiovascular risk is present, SGLT2 inhibitors or GLP 1 receptor agonists may be considered, as they also help reduce visceral fat. Because these drugs can lower lean mass along with fat [21], they should be paired with exercise and enough protein. Lifestyle advice should include resistance exercise, with high intensity interval training where it is safe, to help prevent early sarcopenia [22].

#### Age 65–75 years (transitional sarcopenia)

The glucose sink is shrinking and post meal spikes become more obvious. Sulfonylureas (for example glimepiride) should be reduced or stopped where possible. They push insulin release whatever the glucose level, which raises the risk of hypoglycemia, and extra insulin may not solve a problem that lies in reduced muscle uptake. DPP 4 inhibitors such as teneligliptin or sitagliptin are a reasonable choice, because they lower post meal glucose in a glucose dependent way, rarely cause hypoglycemia and do not cause weight loss that could worsen muscle wasting [19,20]. Progressive resistance training (for example, resistance bands or light weights) should be prescribed to help rebuild muscle [22].

#### Age above 75 years, or frail elderly (SARC F ≥**4)**

In severe sarcopenia, large excursions (ΔBG >150 mg/dL) may reflect a near complete loss of muscle uptake after meals. The main aims are to prevent hypoglycemia and falls.

- *Relax HbA1c targets*. In frail patients who are prone to falls, tight targets (<7.0%) may do more harm than good. A target of about 7.5–8.0% is suggested, and in very poor health the focus should be on avoiding hypoglycemia and symptomatic hyperglycemia [20].
- *Use SGLT2 inhibitors and GLP 1 receptor agonists with caution*. They can cause further weight and lean mass loss in frail patients [21]. They may still be right when there is a strong reason, such as heart failure or chronic kidney disease.
- *Consider alpha glucosidase inhibitors*. Acarbose or voglibose slow carbohydrate absorption in the gut and so reduce the post meal glucose load that weak muscle cannot handle.
- *Lifestyle and diet*. Check that the patient is not relying on a high glycemic “soft diet” (tea and biscuits) because of poor teeth. Offer easily chewable, high quality protein, such as whey, soft paneer or sattu, to limit further muscle breakdown. Resistance exercise should be gentle, supervised where possible, and safe for those at risk of falls.

### 4.2 Limitations

The planned design is cross sectional, so it can describe association but not cause, and reverse causation is possible [18]. Sex, body mass index, HbA1c, diet, physical activity and other illnesses would also need to be recorded. Drug dose, adherence and exact duration of each regimen were not available.

Secondly, SARC F is a self reported screening tool, not a diagnostic test. It is highly specific but has low sensitivity for detecting sarcopenia [23]. Fear of falling, depression or a past fall can raise the score even when muscle mass is normal. Results therefore describe sarcopenia risk and not confirmed sarcopenia. Confirming sarcopenia needs objective tests of muscle mass (appendicular lean mass by DEXA or bioelectrical impedance) and of muscle strength and physical performance, such as handgrip strength and gait speed [8,24]. These tests are costly and hard to arrange in a busy OPD.

Third, in people with diabetes, poor function is not always due to muscle loss. Difficulty climbing stairs or walking may come from diabetic peripheral neuropathy, foot ulcers or knee osteoarthritis, and neuropathy itself is linked to lower muscle strength in T2DM [25].

Exclusion criteria remove only clear cases of non sarcopenic walking difficulty, so some of these patients may remain in the high risk group. Drugs that change body composition, such as SGLT2 inhibitors, could also affect SARC F scores [21].

Lastly, FBS and PPBS are single values taken from existing reports. The timing and size of the meal before PPBS, and the time gap between the glucose test and the SARC F assessment, may vary. Continuous glucose monitoring or a standard meal test would give more reliable measures of the post meal response. Some subgroup estimates were based on few patients, such as only five high risk patients under 65 years.

Future work should be prospective, use objective measures of muscle mass and strength, record drug dose, sex and body composition, and test whether resistance training or a change in drug choice lowers postprandial excursions in patients with a high SARC F score.

## 5. Conclusion

This paper sets out a clear plan for testing whether sarcopenia risk, measured with SARC F, is linked to larger postprandial glycemic excursions in older adults with T2DM and controlled fasting glucose. Implementing rapid functional muscle screening (SARC F) in routine clinical workflows will allow doctors to phenotype their patients accurately. By shifting away from aggressively escalating secretagogues and moving towards age adjusted pharmacology and targeted resistance training, clinicians can achieve safer and more comprehensive glycemic control

## Data Availability

All data produced in the present study are available upon reasonable request to the authors

